## Supporting Information for "Social inequalities, length of hospital stay for chronic conditions and the mediating role of comorbidity and discharge destination: A multilevel analysis of hospital administrative data linked to the population census in Switzerland"

***Table S1** Cluster sizes and Intra-Class-Correlation (ICC) of cluster Variables, linear CCMM (null-model with outcome length of stay)*

| Custer Variable | Records (N) | Clusters (N) | Cluster Size | | | | | ICC |
|---|---|---|---|---|---|---|---|---|
| | | | Min. | Max. | Median | Mean | SD | |
| Hospitals | 141’307 | 188 | 1 | 5513 | 321.50 | 751.63 | 1085.96 | 0.281 |
| Patients | 141’307 | 92’623 | 1 | 23 | 1.00 | 1.53 | 1.02 | 0.154 |

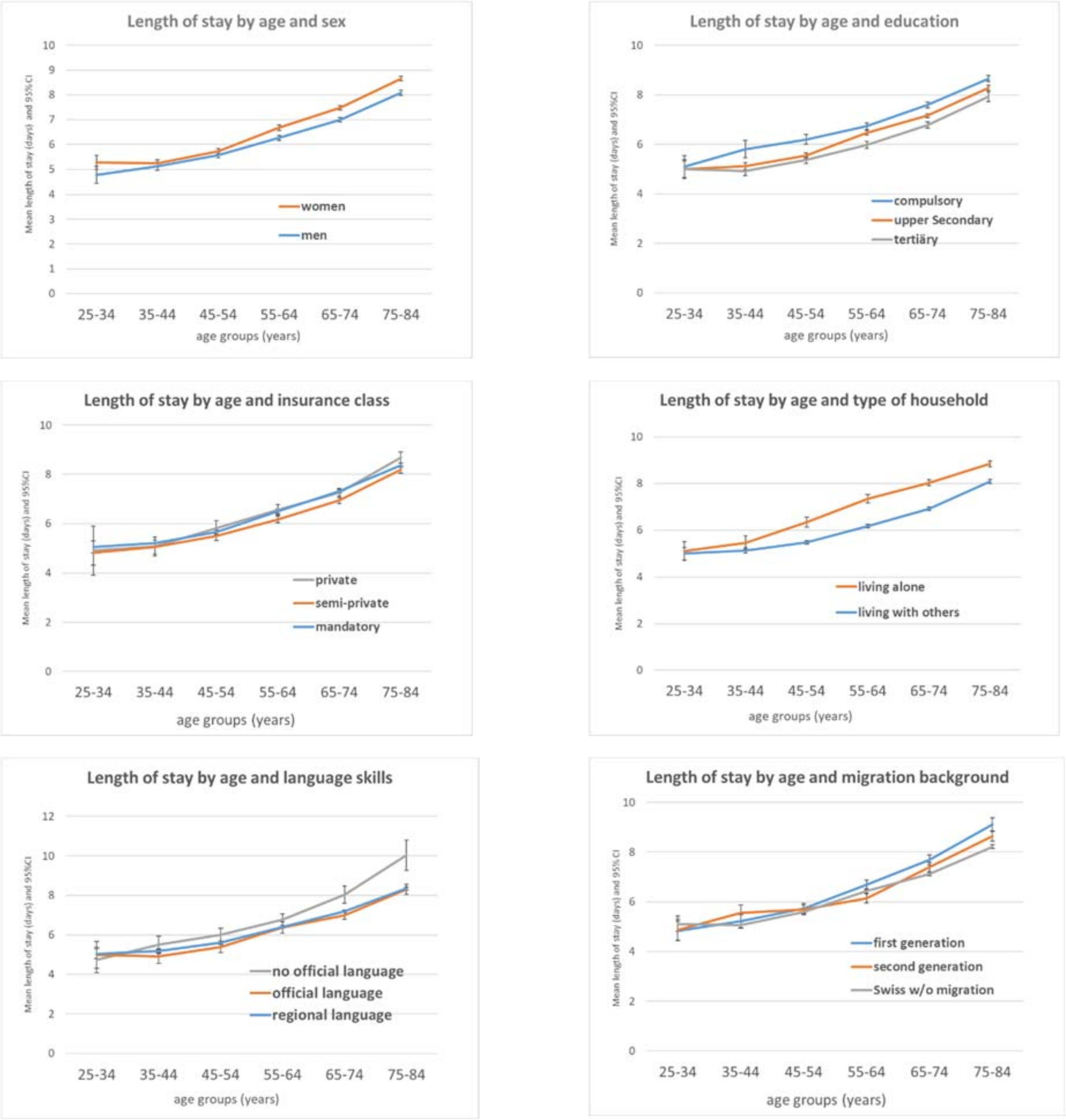


***Figure S1:** Length of stay by age and social factors (top left: sex; top right: education; middle left: insurance class, middle right: type of household; bottom right: language skills; bottom left: migration background*

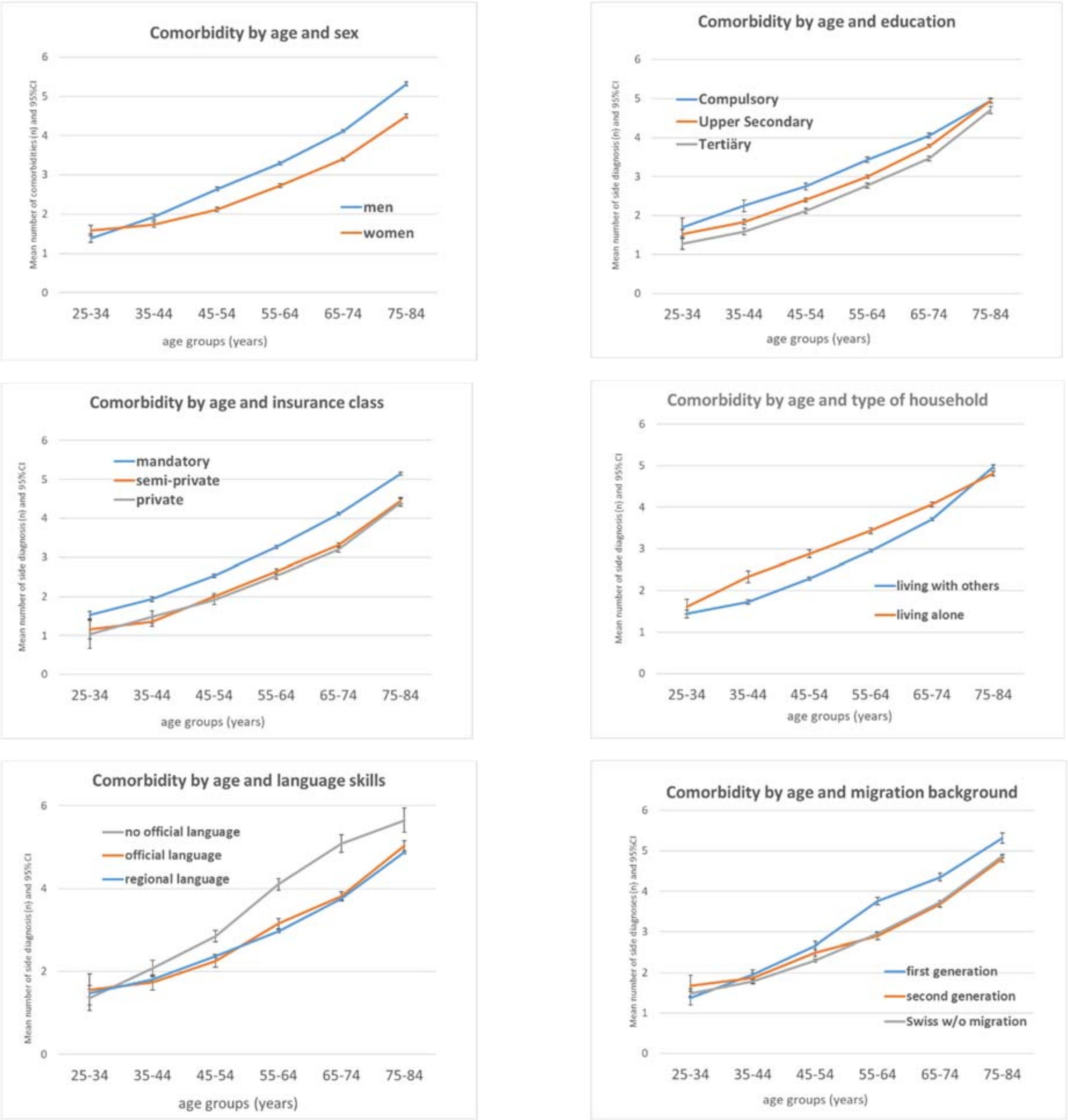


***Figure S2:*** *Comorbidity by age and social factors (top left: sex; top right: education; middle left: insurance class, middle right: type of household; bottom right: language skills; bottom left: migration background*

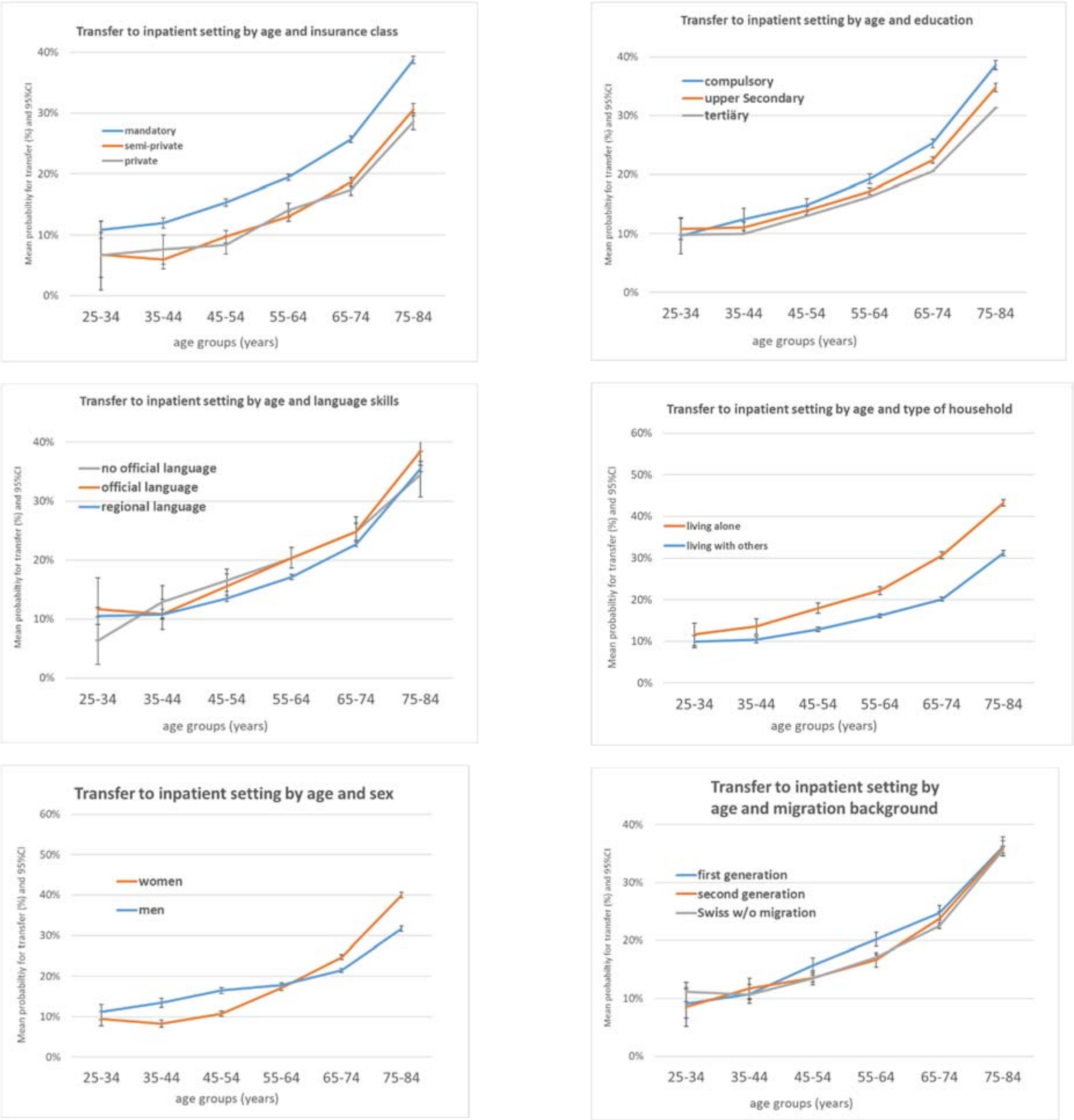


***Figure S3:*** *Discharge destination by age and social factors (top left: sex; top right: education; middle left: insurance class, middle right: type of household; bottom right: language skills; bottom left: migration background*

***Table S2:*** *Effect estimates for significant interactions between main diagnosis and social factors*

| **Simple Contrast for significant interactions between main diagnosis and social factors** | | **Contrast Estimate** | **Adj. Sig.** | **95% Confidence Interval** | |
|---|---|---|---|---|---|
| **Main Diagnosis** | **Educational Attainment** | | | **Lower** | **Upper** |
| Other main diagnoses[1)] | Mandatory vs Tertiary | 0.05 | 0.29 | -0.04 | 0.14 |
| | Upper Secondary vs Tertiary | 0.04 | 0.38 | -0.04 | 0.12 |
| Colon Cancer | Mandatory vs Tertiary | 0.99 | 0.02 | 0.13 | 1.84 |
| COPD | Upper Secondary vs Tertiary | 0.56 | 0.04 | 0.02 | 1.10 |
| Asthma | Mandatory vs Tertiary | 0.93 | 0.04 | 0.06 | 1.80 |
| Ischaemic HD | Upper Secondary vs Tertiary | -0.17 | 0.04 | -0.33 | -0.01 |
| **Main Diagnosis** | **Hospital insurance class** | | | | |
| Other main diagnoses[1)] | private vs. general | 0.36 | <0.001 | 0.18 | 0.53 |
| | semi-private vs general | 0.15 | 0.03 | 0.02 | 0.29 |
| Colon Cancer | semi-private vs general | -1.10 | 0.00 | -1.85 | -0.35 |
| Breast Cancer | private vs. general | -0.56 | 0.01 | -0.99 | -0.13 |
| | semi-private vs general | -0.36 | 0.03 | -0.67 | -0.04 |
| AMI | private vs. general | 0.65 | 0.01 | 0.20 | 1.10 |
| | semi-private vs general | 0.54 | 0.00 | 0.20 | 0.87 |
| COPD | private vs. general | 1.50 | 0.00 | 0.49 | 2.50 |
| | semi-private vs general | 0.92 | 0.01 | 0.27 | 1.58 |
| Back problems | private vs. general | 0.95 | 0.00 | 0.46 | 1.44 |
| | semi-private vs general | 0.61 | 0.00 | 0.28 | 0.93 |
| **Main diagnosis** | **Type of household** | | | | |
| Other main diagnoses[1)] | living alone vs living with others | 0.28 | <0.001 | 0.20 | 0.37 |
| Lung cancer | living alone vs living with others | 0.92 | 0.000 | 0.46 | 1.38 |
| Colon Cancer | living alone vs living with others | 1.21 | 0.001 | 0.49 | 1.94 |
| Back problems | living alone vs living with others | 0.56 | 0.000 | 0.30 | 0.82 |
| **Main diagnosis** | **Languag skills** | | | | |
| Other main diagnoses[2)] | allophone vs. regional language | 0.34 | <0.001 | 0.13 | 0.55 |
| | no regional vs. regional language | 0.00 | 0.98 | -0.12 | 0.13 |
| Colon cancer | allophone vs. regional language | -1.559 | 0.06 | -3.16 | 0.04 |
| COPD | allophone vs. regional language | -0.749 | 0.06 | -1.54 | 0.04 |
| Ischaemic HD | no regional vs. regional language | 0.345 | 0.01 | 0.10 | 0.59 |
| **Main diagnosis** | **Migration background** | | | | |
| Other main diagnoses[2)] | 1st Generation vs Swiss w/o migr.background | 0.16 | <0.001 | 0.05 | 0.27 |
| | 2nd Generation vs Swiss w/o mig. background | 0.02 | 0.68 | -0.07 | 0.10 |
| Colon Cancer | 2nd Generation vs Swiss w/o mig. background | -1.284 | 0.005 | -2.167 | -0.402 |
| Ostheoarthritis | 1st Generation vs Swiss w/o migr.background | 0.019 | 0.838 | -0.163 | 0.201 |
| Back problems | 2nd Generation vs Swiss w/o mig. background | 0.387 | 0.011 | 0.089 | 0.685 |
| The least significant difference adjusted significance level is .05. | | | | | |
| [1)] cf tabel 4; [2)] cf tabel 5 | | | | | |

***Table S3:*** *Associations of comorbidity with social factors (Linear CCMM)*

| **Outcome:** N° of side diagnosis+ | **Modell Comorbidity** (N=140'903) | | | |
|---|---|---|---|---|
| | β (N) | p-value | 95% CI | |
| **Fixed Effects*** | | | Lower | Upper |
| Intercept | 2.70 | 0.109 | -0.60 | 6.00 |
| Educational attainement | | | | |
| Compulsory | 0.37 | 0.000 | 0.33 | 0.41 |
| Upper secondary | 0.23 | 0.000 | 0.19 | 0.26 |
| Tertiary (Ref.) | $0^{b}$ | | | |
| Insurance Class | | | | |
| Private | 0.01 | 0.863 | -0.06 | 0.07 |
| Semi-private | -0.05 | 0.115 | -0.11 | 0.01 |
| Mandatory (Ref.) | $0^{b}$ | | | |
| Household type | | | | |
| Living alone | 0.22 | 0.000 | 0.17 | 0.26 |
| Living with others (Ref.) | $0^{b}$ | | | |
| Sex | | | | |
| Men | 0.23 | 0.000 | 0.18 | 0.27 |
| Women (Ref.) | $0^{b}$ | | | |
| Nationality | | | | |
| Other nationality | 0.32 | 0.000 | 0.21 | 0.42 |
| EU/EFTA | 0.02 | 0.507 | -0.04 | 0.08 |
| Swiss | $0^{b}$ | | | |

+ centred by chronic conditions
*Controlling for clustering on hospital- and patient-level and adjusted for age, chronic condition, language region of hospital and year of discharge

β: Difference in average length of hospital stay to the respective reference category estimated from the mixed linear regression model containing all variables listed in the table and random effects for hospitals and patients.

***Table S4:*** *Associations of discharge destination with social factors and factors related to hospital stay (Logistic CCMM)*

| **Binary Outcome:** Transfer to inpatient setting (vs. discharge to home) | **Model Discharge Destination (N=140'903)** | | | | | | |
|---|---|---|---|---|---|---|---|
| | β (N) | 95% CI | | OR | p-value | 95% CI | |
| **Fixed Effects*** | | Lower | Upper | | | Lower | Upper |
| Intercept | -1.86 | -6.66 | 2.95 | 0.16 | 0.449 | 0.00 | 19.11 |
| Educational attainement | | | | | | | |
| Compulsory | 0.01 | -0.05 | 0.07 | 1.01 | 0.708 | 0.95 | 1.07 |
| Upper secondary | 0.00 | -0.05 | 0.05 | 1.00 | 0.973 | 0.95 | 1.05 |
| Tertiary (Ref.) | $0^{b}$ | | | | | | |
| Insurance Class | | | | | | | |
| Private | -0.08 | -0.21 | 0.04 | 0.92 | 0.196 | 0.81 | 1.04 |
| Semi-private | -0.10 | -0.20 | -0.01 | 0.90 | 0.030 | 0.82 | 0.99 |
| Mandatory (Ref.) | $0^{b}$ | | | | | | |
| Household type | | | | | | | |
| Living alone | 0.45 | 0.40 | 0.50 | 1.56 | 0.000 | 1.49 | 1.64 |
| Living with others (Ref.) | $0^{b}$ | | | | | | |
| Sex | | | | | | | |
| Men | -0.29 | -0.33 | -0.25 | 0.75 | 0.000 | 0.72 | 0.78 |
| Women (Ref.) | $0^{b}$ | | | | | | |
| Nationality | | | | | | | |
| Other nationality | 0.09 | -0.02 | 0.20 | 1.09 | 0.116 | 0.98 | 1.22 |
| EU/EFTA | 0.05 | 0.00 | 0.10 | 1.05 | 0.046 | 1.00 | 1.10 |
| Swiss | $0^{b}$ | | | | | | |
| Comorbidity | | | | | | | |
| NSD (centered by CC) | 0.11 | 0.10 | 0.13 | 1.12 | 0.000 | 1.10 | 1.14 |
| Psychic comorbidity: yes | 0.34 | 0.28 | 0.40 | 1.40 | 0.000 | 1.33 | 1.49 |
| Psychic comorbidity: no (Ref.) | $0^{b}$ | | | | | | |
| Hospital Ward | | | | | | | |
| Surgical | 0.29 | 0.09 | 0.49 | 1.34 | 0.004 | 1.10 | 1.64 |
| Internal medicine or other (Ref.) | $0^{b}$ | | | | | | |
| Need of intensive care | | | | | | | |
| Yes | 0.68 | 0.47 | 0.90 | 1.98 | 0.000 | 1.60 | 2.45 |
| No (Ref.) | $0^{b}$ | | | | | | |

*Controlling for clustering on hospital- and patient-level and adjusted for age, chronic condition, language region of hospital and year of discharge

β: Difference in average length of hospital stay to the respective reference category estimated from the mixed linear regression model containing all variables listed in the table and random effects for hospitals and patients.

***Table S5** Association of length of stay with demographic factors **(age implemented as categorical variable),** social factors, health status and factors related to hospital stay (linear CCMM D)*

| **Outcome: LOS (days)** | **Model D - age categorical** (N=140'903) | | | |
|---|---|---|---|---|
| **Fixed Effects*** | β (days) | p-value | 95% CI Lower | Upper |
| Intercept | 2.25 | <0.001 | 1.34 | 3.15 |
| Educational attainement | | | | |
| Compulsory | 0.05 | 0.25 | -0.04 | 0.14 |
| Upper secondary | 0.04 | 0.35 | -0.04 | 0.12 |
| Tertiary | Ref. | | | |
| Insurance Class | | | | |
| Private | 0.36 | <0.001 | 0.18 | 0.54 |
| Semi-private | 0.16 | 0.02 | 0.02 | 0.29 |
| Mandatory | Ref. | | | |
| Household type | | | | |
| Living alone | 0.28 | <0.001 | 0.20 | 0.37 |
| Living with others | Ref. | | | |
| Sex | | | | |
| Men | -0.30 | <0.001 | -0.38 | -0.22 |
| Women | Ref. | | | |
| Age | | | | |
| 75-84 | 0.67 | <0.001 | 0.31 | 1.03 |
| 65-74 | 0.55 | <0.001 | 0.23 | 0.86 |
| 55-64 | 0.47 | <0.001 | 0.23 | 0.71 |
| 45-54 | 0.27 | <0.001 | 0.09 | 0.46 |
| 25-44 | Ref. | | | |
| Nationality | | | | |
| Other nationality | 0.21 | 0.04 | 0.01 | 0.41 |
| EU/EFTA | 0.14 | 0.02 | 0.02 | 0.26 |
| Swiss | Ref. | | | |
| Chronic Condition (CC) | | | | |
| Lung cancer | 4.81 | <0.001 | 4.38 | 5.24 |
| Colon cancer | 6.97 | <0.001 | 6.27 | 7.67 |
| Breast cancer | 1.53 | <0.001 | 1.17 | 1.89 |
| Prostata cancer | 2.54 | <0.001 | 1.97 | 3.11 |
| Diabetes w/o complication | 4.03 | <0.001 | 3.04 | 5.02 |
| Diabetes with complication | 7.49 | <0.001 | 6.79 | 8.19 |
| Acute myocardial infarction | 0.70 | <0.001 | 0.39 | 1.02 |
| Acute cerebrovascular diseases | 3.92 | <0.001 | 3.41 | 4.43 |
| Congestive heart failure | 6.41 | <0.001 | 5.93 | 6.90 |
| COPD | 5.09 | <0.001 | 4.61 | 5.57 |
| Asthma | 2.00 | <0.001 | 1.63 | 2.37 |
| Ostheoarthritis | 2.93 | <0.001 | 2.38 | 3.49 |
| Back problems | 3.50 | <0.001 | 3.00 | 3.99 |
| Disc disorder | 2.51 | <0.001 | 1.93 | 3.09 |
| Ischämic heart disease | Ref. | | | |
| Comorbidity | | | | |
| NSD (centered by CC) | 0.80 | <0.001 | 0.70 | 0.89 |
| Psychic comorbidity: yes | 0.36 | <0.001 | 0.14 | 0.57 |
| Psychic comorbidity: no | Ref. | | | |
| Hospital Ward | | | | |
| Surgical | 1.72 | <0.001 | 1.24 | 2.21 |
| Internal medicine or other | Ref. | | | |
| Need of intensive care | | | | |
| Yes | 3.15 | <0.001 | 2.78 | 3.53 |
| No | Ref. | | | |
| Discharge destination | | | | |
| Died in hospital | 0.34 | 0.31 | -0.32 | 1.01 |
| Transfer to inpatient setting | 1.99 | 0.00 | 1.76 | 2.22 |
| Discharge to home | Ref. | | | |
| Akaike criterion, corrected | 868'829 | | | |

*The model controls for clustering on hospital- and on patient-level and is adjusted for language region of hospital and year of discharge

The regression coefficients β are the estimated differences in average length of stay between the respective category and the reference category obtained from the respective model.
